# Evaluating the Etiology of Metallic Taste During Head and Neck Cancer Treatments: A Study of Facial and Glossopharyngeal Nerve Interactions

**DOI:** 10.1101/2024.11.02.24316658

**Authors:** Guillaume Buiret, Brignot Hélène, Septier Chantal, Feron Gilles, Thomas-Danguin Thierry

## Abstract

Metallic Taste (MT) is frequently described during head and neck cancer treatments but very little is known about its etiologies. One hypothesis to explain the MT is the removal of facial nerve inhibition on the glossopharyngeal nerve. Indeed, the decrease of taste afferents mediated by the facial nerve (anterior two-thirds of the tongue) due to cancer or its treatments, would reveal those mediated by the glossopharyngeal nerve (posterior one-third of the tongue) and thus lead to MT perception. The aim of this study was to evaluate the validity of this hypothesis.

Selective supraliminar taste tests on the tip and the base of the tongue were regularly performed on 44 patients with head and neck cancers before, during, and after their treatment. Sweet, salty, bitter, sour, and MT were tested. Patients were grouped based on whether they reported experiencing MT or not.

12 patients complained about MT (27.2%), always during the treatment phase. Most of them (83.3%) were treated by surgery and radiotherapy or radiochemotherapy. Supraliminar tastes were altered in every patient, especially during the treatment phase. Test results showed that perceived intensity was significantly reduced in patients reporting MT for salt, sweet and sour. This was observed more on the base of tongue than on the tip of the tongue. MT was significantly linked with mucositis (p=0.027) but with neither candidiasis (p=0.38) nor salivary flow (p=0.63).

The hypothesis of removal of facial nerve inhibition on the glossopharyngeal nerve cannot explain MT in head and neck cancer.

## INTRODUCTION

Head and Neck Cancers (HNC), encompassing a diverse range of malignant tumors affecting anatomical structures crucial for speech, swallowing, and breathing, present a substantial challenge in the field of oncology. First, even if the incidence is decreasing, they are the fifth most common cancers in men worldwide (1). HNC are characterized by their significant impact on patients’ quality of life due to symptoms that impair essential oral and throat functions (2-4). Among the numerous manifestations associated with these cancers, Metallic Taste (MT) stands out as one of the most complex and poorly understood symptoms. MT is described by patients as a persistent sensation of metallic flavor in the mouth. It can manifest at various stages of the disease, whether during diagnosis, treatment, or remission. This disruption of the sense of taste has profound consequences on appetite, nutrition, and quality of life of patients and can even influence their adherence to treatment, as it may contribute to weight loss and malnutrition (5-7). While MT is widely acknowledged by healthcare professionals as a common side effect of cancers in general and Head and Neck Cancers (HNC) particularly (8, 9), its understanding, management, and underlying mechanisms remain inadequately explored. This knowledge gap hinders our ability to offer appropriate treatment strategies to alleviate this symptomatic manifestation.

According to Logan et al. (10) or Reith et al. (11), the causes of MT are not understood. It may be linked with oral health issues (mucositis, candidiasis), a neuropathy, the reduction in taste receptors, a direct stimulation of a specific MT receptor close to bitter receptors (T2R) or receptors of the TRPV1 family, the toxicity to salivary glands, the intra-oral lipoperoxidation. Among the possible causes, the removal of facial nerve inhibition on the glossopharyngeal nerve may be hypothesized (12). Under normal circumstances, the facial nerve may exert an inhibitory effect on the glossopharyngeal nerve to regulate taste sensation. Because of the cancer and/or its treatments, the decrease of taste afferents mediated by the facial nerve (anterior two-thirds of the tongue) would reveal those mediated by the glossopharyngeal nerve (posterior one-third of the tongue), leading to abnormal taste sensations such as MT. This lack of inhibition effect is commonly evoked after middle ear surgery (12-16) but has never been proven in HNC.

In case of release of inhibition of the facial nerve on the glossopharyngeal nerve, the following results are expected: for one or more tastes, the taste on the tongue base should be significantly more intense than on the tip of the tongue in case of MT. In this study, we tested this hypothesis by performing supraliminar taste tests in both territories of the taste nerves on the tongue in patients with HNC, regardless of their treatments. The perceived intensity of sweet, salty, bitter, sour, metallic tastes was recorded following the application of specific taste-eliciting compounds on the anterior and posterior parts of the tongue, as well as on both sides (left and right). Since MT could occur at various stages of the treatment protocol, each patient was tested at multiple time points before, during, and after treatment.

## MATERIAL AND METHODS

### Study population

The prospective study involved a cohort of 44 patients diagnosed with HNC. These patients were included at the onset of their cancer, before the occurrence of MT and were followed-up during one year, unless a recurrence or another cancer occurs.

### Supraliminar recognition then intensity taste testing

Selective supraliminar taste tests were conducted on each patient at regular intervals. Identical tests were performed before any treatment ((m) moment 1); after surgery if any (m2); in the middle (m3) and at the end (m4) of the radiotherapy if any; at 3 (m5)-6 (m6)-9 (m7) and 12 (m8) months. Taste assessments were carried out on four distinct regions of the tongue: the tip (taste mediated by the facial nerve) and the base (taste mediated by the glossopharyngeal nerve) and each on the right and on the left. Figure S1 in the appendix presents the test locations on the tongue. A cotton swab, soaked in a solution of tastant diluted in Evian water, was applied to each of the predefined lingual locations (tip/base, left/right) following a sequence that avoided anticipation through iterative testing. The evaluation encompassed the following tastes (tastants): sweet (sucrose, 850g/l), salty (sodium chloride, 75g/l), bitter (anhydrous caffeine, 20g/l), sour (citric acid, 50g/l), and metallic (iron sulfate, 10 g/l). All tastants were purchased at Merck (Saint-Quentin-Fallavier, France). Each patient was asked first to identify the taste and then to rate the perceived intensity on a numerical scale from 0 (no perception) to 5 (very intense) at each of the four predefined lingual locations. The mouth was rinsed with Evian water between each test.

### Other tests

The oral mucositis and candidiasis were determined by oral examination by an ENT (0 if no, 1 if yes). The non-stimulated salivary flow was determined by weighing the saliva collected over a 10-minute period as described previously (15).

### Data Analysis

At the end of the study, patients were divided into two groups based on whether they reported experiencing metallic taste (MT) or not.

Confidence intervals by moments and tongue locations were calculated for different time points and tongue locations to compare taste intensity between patients who reported experiencing MT and those who did not. According to Neyman and Pearson (17), if the confidence intervals don’t cross on the graph, the test was considered as significant.

Links between oral mucositis and MT status and candidiasis status and MT status were determined by chi-squared tests.

The potential link between salivary flow reduction and MT was determined by comparing the mean flow according to MT status by a t-test.

A p-value of <0.05 was considered statistically significant for every test in this study.

### Ethical Considerations

This study was conducted in accordance with the French ethical guidelines and principles related to biomedical research. An informed consent was obtained from all participating patients. The whole study received approval from the ethic committee CPP Est I, Dijon, France (2017-A03641-52). The protocol was registered on Clinicaltrials.gov (NCT03558789).

## RESULTS

44 patients were recruited and followed up for one year. The flowchart is reported in figure 1. The characteristics of the population are presented in table 1. As in every study on HNC, the patients were predominantly male, middle-aged, and with advanced Tumor and Node stages. The treatment was predominantly multimodal.

**Table 1.** population characteristics

|  |  |
| --- | --- |
| Sex: males n(%) ; females n(%) | 34 (77.3%), 10 (22.7%) |
| Mean age at diagnosis ± SD (year) | 63.1 ± 9.8 |
| Location n (%):<br>Hypopharynx<br>Larynx<br>Oropharynx<br>Oral Cavity<br>Unknown primary | 3 (6.8%)<br>8 (18.2%)<br>14 (31.8%)<br>14 (31.8%)<br>5 (11.4%) |
| Tumor stage n (%):<br>1<br>2<br>3<br>4<br>x | 9 (20.4%)<br>18 (40.9%)<br>8 (18.2%)<br>4 (9.1%)<br>5 (11.4%) |
| Node stage n (%):<br>0<br>1<br>2a<br>2b<br>2c<br>3a | 20 (45.5%)<br>10 (22.7%)<br>4 (9.1%)<br>5 (11.4%)<br>2 (4.5%)<br>3 (6.8%) |
| Smoking at inclusion n (%):<br>Yes<br>No<br>Quit | 20 (45.5%)<br>5 (11.4%)<br>19 (43.1%) |
| Treatment<br>Withdrawal of consent before treatment<br>Chemotherapy, surgery, radiotherapy<br>Surgery<br>Surgery, radiotherapy<br>Surgery, radiochemotherapy | 5 (11.4%)<br>1 (2.2%)<br>14 (31.8%)<br>12 (27.3%)<br>12 (27.3%) |

**Figure 1.**
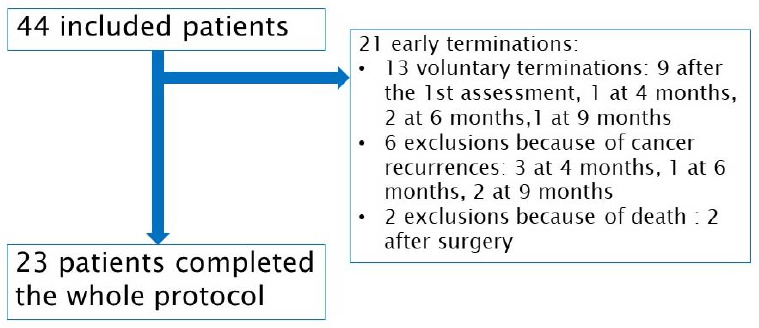
study flowchart

Among the 23 patients who completed the whole study, 12 complained of MT (27.2%) ; they all reported MT during the treatment phase (moments 2 to 5, one patient still at moment 6). Ten of these patients (83.3%) were treated by surgery and radiotherapy or radiochemotherapy, and two others (16.7%) by surgery only.

The figures 2 to 5 represent the evolution of mean taste intensity as a function of the time of treatment, and according to the reported MT status for the bitter, salty, sour, sweet respectively. The figure S2 in the appendix represent the same results for the metallic taste. The results of all chi-2 tests to evaluate differences between groups of patients for taste recognition and t-tests for taste intensity are presented in the supplement (table S1 and S2) respectively. No differences were observed for taste recognition. Most of the intensities were not significantly different in function of MT status and tongue location. Nevertheless, intensities were significantly reduced in patients with metallic taste for salty (figure 3), sour (figure 4) and sweet (figure 5) tastes, during treatments (most of the time at the end of radio(chemo)therapy, i.e. moment 4). The differences were observed on the base of tongue locations as well as on the tip of the tongue.

**Figure 2.**
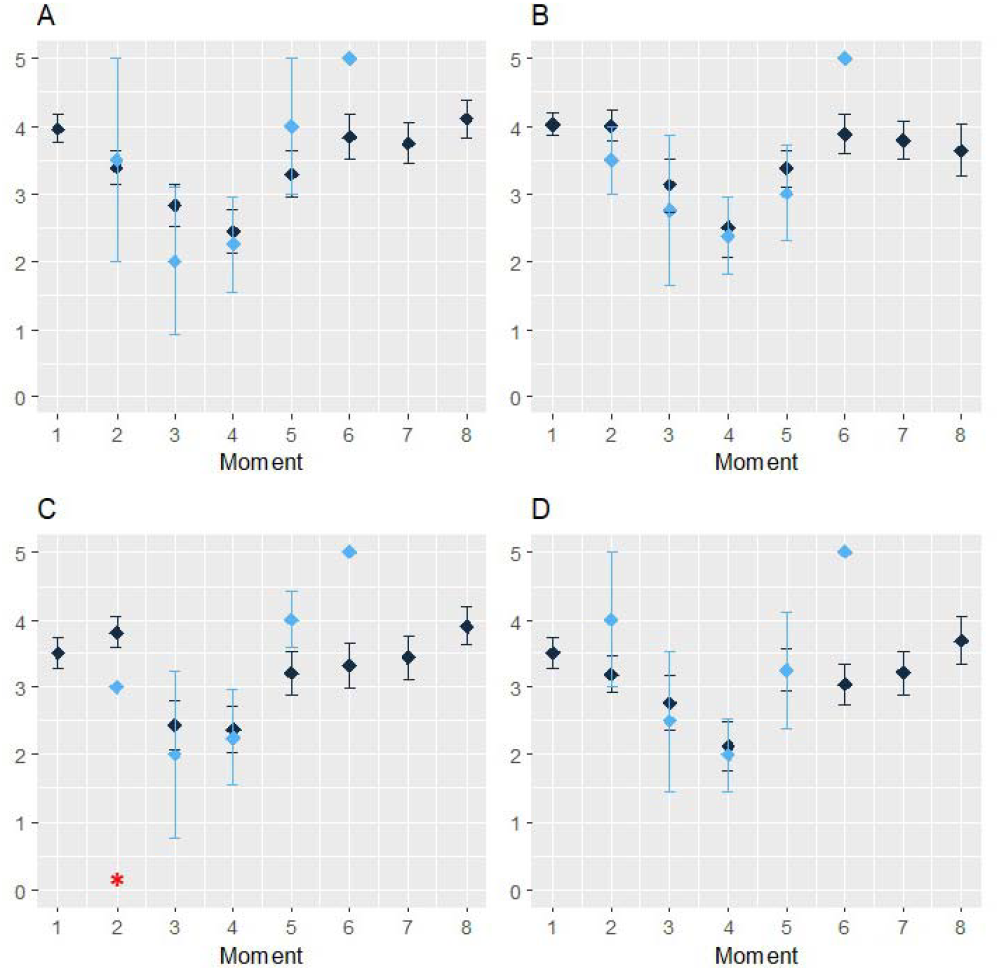
evolution of results for bitter taste according to the precise location on the tongue Blue = metallic taste, black = no metallic taste. A = right tongue base, B = left tongue base, C = left tip of the tongue, D = right tip of the tongue Red *: significancy of t-test<0.05 Moments: (m) moment 1); after surgery if any (m2); in the middle (m3) and at the end (m4) of the radiotherapy if any; at 3 (m5)-6 (m6)-9 (m7) and 12 (m8) months.

**Figure 3.**
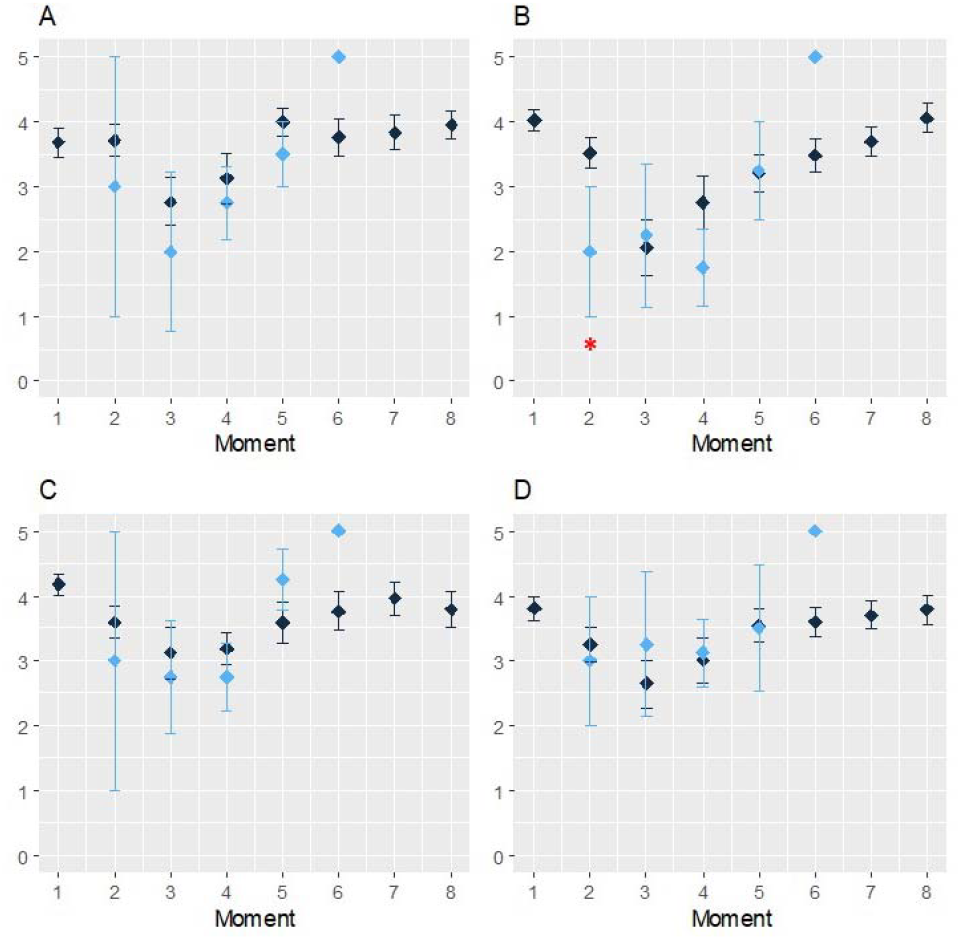
evolution of taste results for salty taste according to the precise location on the tongue Blue = metallic taste, black = no metallic taste. A = right tongue base, B = left tongue base, C = left tip of the tongue, D = right tip of the tongue Red *: significancy of t-test<0.05 Moments: (m) moment 1); after surgery if any (m2); in the middle (m3) and at the end (m4) of the radiotherapy if any; at 3 (m5)-6 (m6)-9 (m7) and 12 (m8) months.

**Figure 4.**
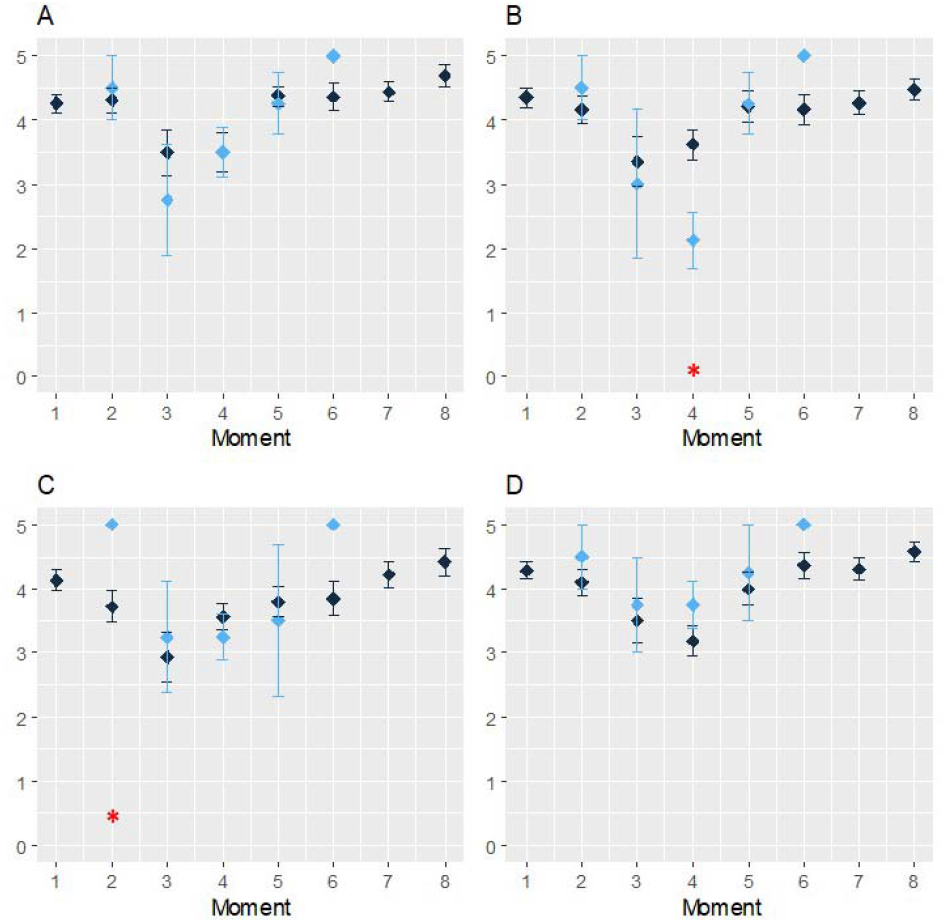
evolution of taste results for sour according to the precise location on the tongue Blue = metallic taste, black = no metallic taste. A = right tongue base, B = left tongue base, C = left tip of the tongue, D = right tip of the tongue Red *: significancy of t-test<0.05 Moments: (m) moment 1); after surgery if any (m2); in the middle (m3) and at the end (m4) of the radiotherapy if any; at 3 (m5)-6 (m6)-9 (m7) and 12 (m8) months.

**Figure 5.**
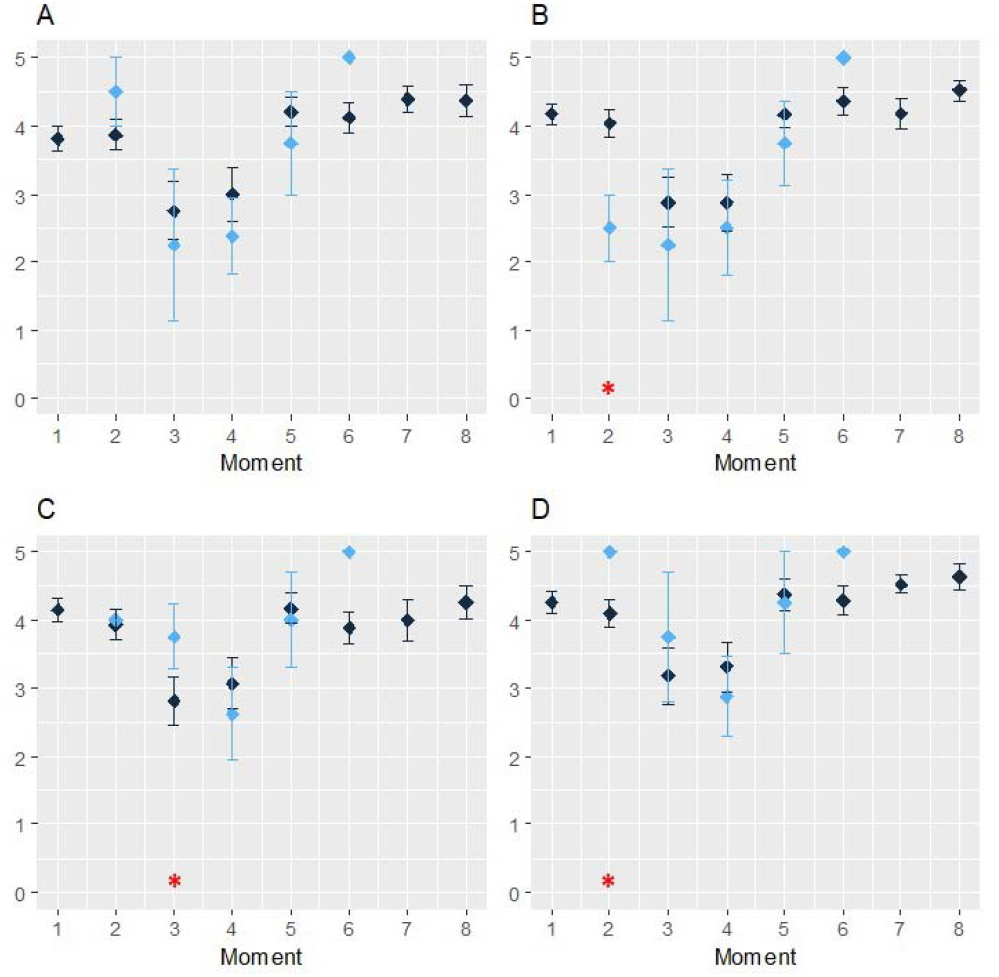
evolution of taste results for sweet according to the precise location on the tongue Blue = metallic taste, black = no metallic taste. A = right tongue base, B = left tongue base, C = left tip of the tongue, D = right tip of the tongue Red *: significancy of t-test<0.05 Moments: (m) moment 1); after surgery if any (m2); in the middle (m3) and at the end (m4) of the radiotherapy if any; at 3 (m5)-6 (m6)-9 (m7) and 12 (m8) months.

There was only one patient who reported MT at moment 6 so no confidence interval was made possible and statistical significancy was not retained.

**Table 2:** intensity differences (posterior minus anterior) of tastes according to metallic taste (MT) status

|  | patients reporting MT | patients reporting No MT | p-value |
| --- | --- | --- | --- |
| Bitter | 0.2 ± 1.5 | 0.7 ± 1.9 | 0.177 |
| Salty | -1.3 ± 2.1 | -0.1 ± 1.7 | 0.031 |
| Sour | -0.7 ± 2.0 | 0.4 ± 1.6 | 0.028 |
| Sweet | -1.3 ± 2.3 | -0.2 ± 1.4 | 0.056 |

For each taste, intensities recorded when applying stimuli on the tip of the tongue (i.e. facial nerve mediation) and on the tongue base (i.e. glossopharyngeal nerve mediation) were added up. The mean differences (posterior minus anterior) were then compared according to the MT status. The results are presented in Table 2.

For salty and sour tastes, the mean intensity was significantly lower on the tongue base than on the tip of the tongue in the case of patients reporting MT (p=0.031 and 0.028 respectively). This was almost significant for sweet taste (p=0.056).

A negative value means that the intensity was higher on the tip of the tongue than on the tongue base. A positive value mean means that the intensity was higher on the tongue base than on the tip of the tongue.

To summarize, salty and sour tastes intensities on the tip of the tongue were lower than on the tongue base for patients reporting MT compared to patients who did not report MT. The difference was marginally significant for sweet taste but not significant for MT.

Besides, MT was significantly linked with mucositis (p=0.027) but with neither candidiasis (p=0.38) nor with salivary flow (p=0.63).

## DISCUSSION

Our findings reveal that the intensities of all tastes decreased across various regions of the tongue during HNC treatments. Although the decline was more pronounced at the base of the tongue than at the tip, there were no significant differences for most of the tests. This disproves the hypothesis that the removal of facial nerve inhibition on the glossopharyngeal nerve is responsible for this phenomenon (12). Instead, this aligns with Deshpande et al. (18), who suggested that dysgeusia in HNC is more likely due to a reduced density of taste buds rather than nerve damage.

### Changes in Taste Perception During HNC Treatments

The observed pattern of side-effects peaking during irradiation and then recovering, as seen in our study, is commonly reported in other series for taste intensity tests (20-23) and salivary flows (24). Sapir et al. also did not find any correlation between dysgeusia and salivary flow, although they did find a correlation between dysgeusia and xerostomia questionnaire responses (24).

All taste intensities decreased during HNC treatments, with a subsequent recovery. There were some differences between the two groups of patients based on reported MT, regarding salty and sour tastes, possibly due to higher alteration of ion channel receptors. Interestingly, our findings showed an inverse relationship between the expected results for different tongue regions. The base of the tongue, typically less sensitive to these tastes, showed a greater decrease in taste intensity compared to the tip. This could suggest a more complex mechanism involving different receptor types and their distribution across the tongue. Finally, the recovery phase observed in taste intensities mirrors patterns found in other studies (20-23).

### Metallic Taste (MT) and Its Mechanisms

When specifically considering MT, its incidence in our study was 27.2%, which is consistent with other series (9). Previous studies on tympanic cord anesthesia, a branch of the facial nerve involved in taste, have shown a significant increase in bitter intensity at the base of the tongue and MT (13-16). However, our results do not support a potential link between bitter receptors and MT (19), as bitter intensity did not significantly differ between MT and non-MT patients (Figure 2, Tables S1 and S2). This lack of significant differences highlights the need for further research into the specific mechanisms underlying these changes in taste perception during and after HNC treatments. Moreover, MT was significantly associated with mucositis but not with candidiasis or salivary flow. Given that the removal of facial nerve inhibition on the glossopharyngeal nerve seems to be disproved and MT is linked to mucositis but not candidiasis, the most probable explanation for MT is inflammation, specifically lipoperoxidation. Malondialdehyde, a byproduct of lipoperoxidation, has been shown to be elevated in HNC patients (25). Our study included salivary samplings to measure salivary levels of malondialdehyde.

In summary, our study challenges existing hypotheses regarding the cause of MT in HNC patients and suggests that inflammation, rather than nerve damage, may play a significant role. The dynamic changes in taste perception, along with the interplay between different types of taste receptors and regions of the tongue, warrant further investigation. Understanding these mechanisms can lead to more effective interventions for managing dysgeusia in these patients. Furthermore, the complex interplay between taste perception and the altered microenvironment of the oral cavity in HNC patients, including changes in salivary composition and flow, remains a hypothesis that requires testing.

## CONCLUSION

The hypothesis of removal of facial nerve inhibition on the glossopharyngeal nerve cannot explain metallic taste in HNC. MT was significantly linked with mucositis, suggesting a possible role of inflammation, potentially lipoperoxidation.

## Data Availability

All data produced are available online at https://hal.science/hal-04750951

## Aknowledgements

to C.T. Molta, MD, for the English translation.

