## Supplements for "Evaluating the Etiology of Metallic Taste During Head and Neck Cancer Treatments: A Study of Facial and Glossopharyngeal Nerve Interactions"

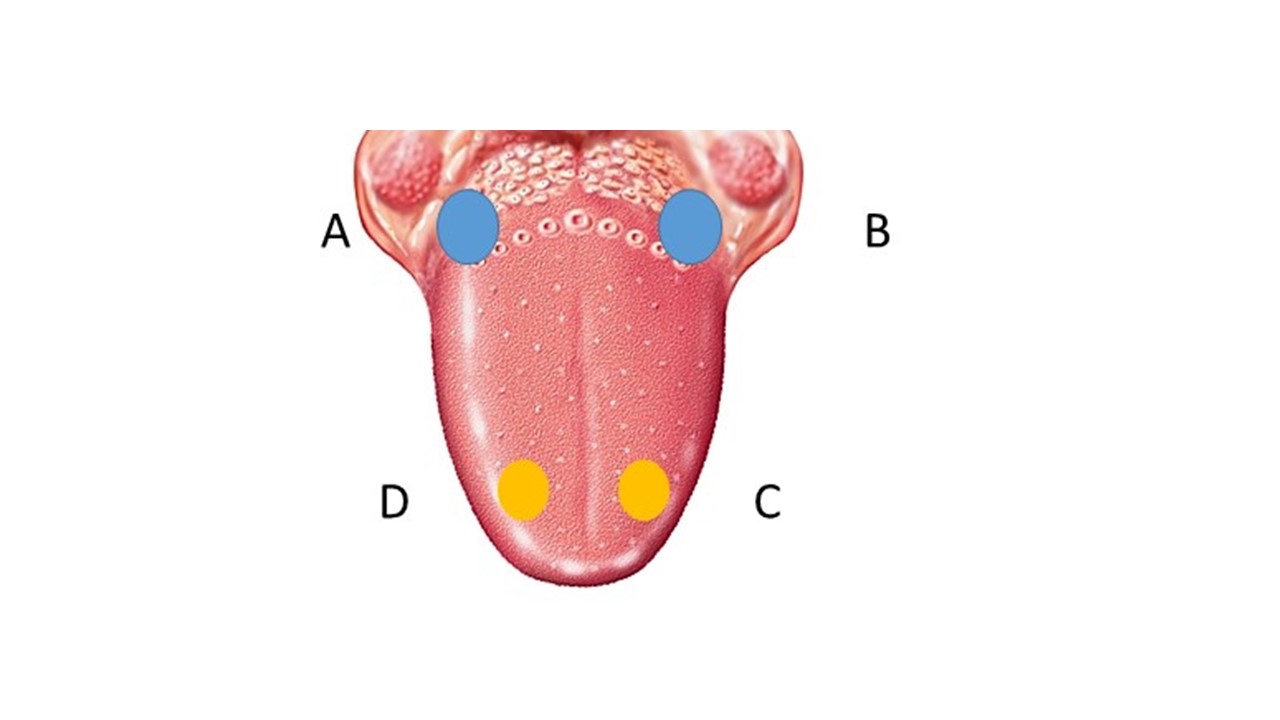

Figure S1: tests location on the tongue

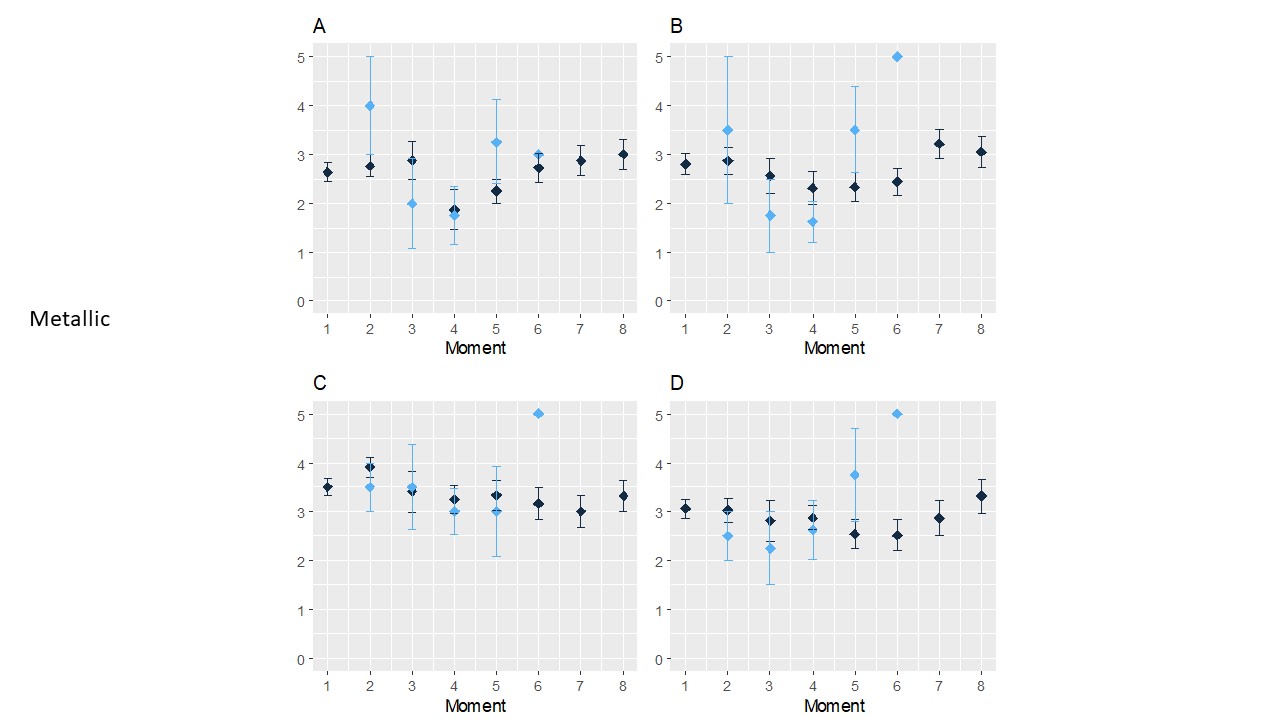

Figure S2: evolution of taste results for metallic according to the precise location on the tongue

Blue = metallic taste, black = no metallic taste. A = right tongue base, B = left tongue base, C = left tip of the tongue, D = right tip of the tongue

Moments: (m) moment 1); after surgery if any (m2); in the middle (m3) and at the end (m4) of the radiotherapy if any; at 3 (m5)-6 (m6)-9 (m7) and 12 (m8) months.

| Taste | Location | Moment 2 | Moment 3 | Moment 4 | Moment 5 |
| --- | --- | --- | --- | --- | --- |
| Water | A | 0.4058 | 0.6524 | 0.3743 | 1 |
|  | B | 0.5489 | 1 | 0.2664 | 1 |
|  | C | 0.4929 | 1 | 0.3743 | 0.4863 |
| Salty | A | 1 | 1 | 0.1111 | 0.3912 |
|  | B | 0.6673 | 0.6604 | 0.8836 | 0.9358 |
|  | C | 0.8188 | 1 | 0.0506 | 1 |
|  | D | 0.4145 | 0.8442 | 1 | 0.1603 |
| Sour | A | 1 | 0.1973 | 0.8783 | 1 |
|  | B | 1 | 1 | 0.4440 | 1 |
|  | C | 1 | 0.8098 | 1 | 0.2679 |
|  | D | 1 | 1 | 1 | 0.6532 |
| Sweet | A | 1 | 0.9067 | 0.6547 | 1 |
|  | B | 1 | 0.7144 | 0.1797 | 0.6542 |
|  | C | 1 | 1 | 0.8738 | 1 |
|  | D | 1 | 0.7109 | 1 | 1 |
| Bitter | A | 1 | 0.8442 | 1 | 1 |
|  | B | 1 | 1 | 0.6171 | 1 |
|  | C | 0.4389 | 1 | 0.4689 | 0.8151 |
|  | D | 0.7535 | 1 | 0.6171 | 1 |
| Metallic | A | 1 | 1 | 1 | 0.3045 |
|  | B | 0.6422 | 0.9067 | 0.2664 | 0.2272 |
|  | C | 1 | 1 | 0.6171 | 1 |
|  | D | 1 | 1 | 0.6650 | 0.8151 |

Tableau S1 : results of chi-square tests of taste recognition according to the moment

| Taste | Location | Moment 2 | Moment 3 | Moment 4 | Moment 5 |
| --- | --- | --- | --- | --- | --- |
| Water | A | 0.6804 | **0.0015** | 0.2842 | 0.6942 |
|  | B | 0.5268 | 0.8089 | **0.0458** | 0.8342 |
|  | C | 0.4929 | 0.7732 | 0.8971 | 0.4178 |
| Salty | A | 0.783 | 0.5858 | 0.5898 | 0.4084 |
|  | B | 0.3613 | 0.8802 | 0.184 | 0.9611 |
|  | C | 0.8165 | 0.7148 | 0.4862 | 0.285 |
|  | D | 0.8450 | 0.6354 | 0.8428 | 0.9687 |
| Sour | A | 0.7601 | 0.4616 | 1 | 0.8173 |
|  | B | 0.6195 | 0.7873 | **0.0120** | 0.9412 |
|  | C | **<10^-5^** | 0.7565 | 0.4705 | 0.8245 |
|  | D | 0.5628 | 0.7766 | 0.2152 | 0.7688 |
| Sweet | A | 0.4043 | 0.7601 | 0.6956 | 0.3810 |
|  | B | **0.0162** | 0.6230 | 0.6578 | 0.5651 |
|  | C | 0.7734 | **0.0412** | 0.5830 | 0.8342 |
|  | D | **<10^-5^** | 0.6059 | 0.8339 | 0.8818 |
| Bitter | A | 0.9523 | 0.5097 | 0.8134 | 0.5423 |
|  | B | 0.4881 | 0.7663 | 0.8645 | 0.6472 |
|  | C | **0.0012** | 0.7510 | 0.8763 | 0.1678 |
|  | D | 0.5621 | 0.8245 | 0.8495 | 1 |
| Metallic | A | 0.4262 | 0.4245 | 0.8641 | 0.3316 |
|  | B | 0.7459 | 0.3774 | 0.2262 | 0.2758 |
|  | C | 0.5604 | 0.9309 | 0.6566 | 0.7479 |
|  | D | 0.4648 | 0.5414 | 0.7082 | 0.2968 |

Tableau S2 : results of t-tests of tastes intensities according to the moment
